# Differentiated HIV Service Delivery vs Conventional Care: Tuberculosis Preventive Therapy Outcomes for People Living with HIV in Sub-Saharan Africa

**DOI:** 10.1101/2025.01.15.25320590

**Authors:** Ann Johnson, Lucy Chimoyi, Salome Charalambous, Nicole Kawaza, Chris J Hoffmann, J. Lucian Davis, Violet Chihota

**Affiliations:** Yale School of Medicine, 333 Cedar St, New Haven, CT 06510, USA; Epidemiology of Microbial Diseases, Yale School of Public Health, 60 College St, New Haven, CT 06510, USA; Implementation Research Division, The Aurum Institute, 29 Queens Rd, Parktown, Johannesburg 2194, South Africa; School of Public Health, University of the Witwatersrand, 60 York Rd, Parktown, Johannesburg 2193, South Africa; School of Medicine, Johns Hopkins, 725 N. Wolfe Street, Baltimore, Maryland, 21205, US; Pulmonary, Critical Care, and Sleep Medicine Section, Department of Internal Medicine, Yale School of Medicine, 333 Cedar St, New Haven, CT 06510, USA

## Abstract

**Introduction:** Differentiated service delivery (DSD) models, which are mechanisms of HIV care that reduce provider visits and offer varied ART delivery methods, are scaling up across sub-Saharan Africa. It is unknown how the movement of patients to DSD models impacts services beyond ART, including the uptake and completion of tuberculosis preventive therapy (TPT).

**Methods:** Using the RE-AIM framework, we analyzed data from Opt4TPT, a longitudinal cohort study examining TPT delivery in South Africa and Zimbabwe. We constructed multivariate logistic regression models to evaluate the association of receiving ART from a DSD model with the proportion of participants who initiated and completed TPT, as measured by electronic medication boxes. We constructed a Cox proportional hazards model to assess the association between DSD models and time to TPT initiation.

**Results:** Among 1193 participants, 276 received ART through a DSD model, while 917 used the conventional model. Overall, 1035 (87%) initiated TPT, including 242 (88%) in DSD models and 793 (86%) in conventional models. Receiving ART from a DSD model was not significantly associated (OR 1.11, 95% CI 0.74-1.67, p = 0.61) with TPT initiation. DSD models had a significantly longer mean time to initiation (6.5 vs. 2.7 days, p = 0.01). Of the 731 (71%) participants with MERM box data, 356 (49%) completed TPT. Bivariate analysis showed significantly higher odds of completing TPT among those in DSD models (OR 1.53, 95% CI 1.06- 2.21, p=0.024). This association was not significant in multivariate analysis after adjusting for demographic and clinical factors (OR 0.89, 95% CI 0.58-1.36, p=0.58).

**Conclusions:** We found high TPT uptake in DSD and conventional models of care, indicating that TPT delivery in DSD models is feasible. We found low TPT completion in both models of care, showing a need to focus on improving TPT completion overall.

## Introduction

Since 2016, the WHO has recommended implementing differentiated service delivery (DSD) HIV care models to reduce the healthcare system burden and expand access to antiretroviral therapy (ART) (1). In contrast to the conventional model for ART delivery, which requires frequent, often monthly, in-clinic visits and provider consultations, Differentiated Service Delivery (DSD) models allow “stable” patients, defined either as an undetectable viral load or a minimum of 12 months on ART (2), to obtain ART refills through community-based pick-ups or fast-track clinic pickups with less frequent provider visits (3). DSD models retain patients in care at similar rates to conventional models while reducing costs for the healthcare system and increasing patient satisfaction through reduced costs and greater personalization of care (4–6). Clinically, DSD models maintain viral suppression levels equivalent to those in conventional models (7–9).

DSD models tailor HIV services, including the location of ART refill, degree of provider support, and frequency of visits, according to individual patient needs and preferences. In sub-Saharan Africa, DSD models are being rapidly scaled up (2). In South Africa, between January 2018 and October 2019, the number of patients actively receiving ART through a DSD model increased from 1.5 million to 2 million (10). In both South Africa (10) and Zimbabwe (11,12) in 2018, around 35% of virally suppressed PLHIV were receiving care through DSD models.

As DSD models continue to scale up, the WHO recommends integrating other essential forms of care, including tuberculosis preventive therapy (TPT) — a crucial treatment to prevent death from tuberculosis (13) — and care for reproductive and non-communicable diseases (14), primarily hypertension and diabetes, with DSD models (15,16). In South Africa in 2019, about 500,000 patients received medications for noncommunicable diseases through DSD models, and in 2021, a low (4%) percentage of the prescriptions in DSD models were for TPT (17). DSD models have demonstrated adaptability, particularly in maintaining continuity of care during the COVID-19 pandemic (18). However, potential barriers threaten the successful expansion of DSD models to these other aspects of care, including whether the infrequent contact with providers in DSD models might limit the ability of PLHIV to access preventive care and care for different health issues (19–21). There is limited evidence evaluating the delivery of TPT and NCD treatments through DSD models. As DSD models continue to scale up, ensuring that PLHIV receiving care from DSD models have access to TPT is essential for reducing TB incidence in PLHIV (22).

Our study aimed to assess the impact of conventional versus differentiated service delivery (DSD) models on the implementation of tuberculosis preventive therapy (TPT) for PLHIV (23). We hypothesized that initiation of TPT in DSD models would be delayed compared to conventional models because the reduction in clinician interactions in DSD models could reduce clinicians’ time for preventive care, such as TPT initiation. Understanding how TPT can be delivered in DSD models can also help improve these models for integrating treatment for non-communicable diseases.

## Methods

### Study Design and Setting

We conducted a secondary analysis of the Opt4TPT study data. This prospective, observational cohort study was a programmatic assessment of the implementation of TPT among PLHIV in three countries in sub-Saharan Africa between July 2021 and December 2023. We evaluated the association between ART delivery models, conventional versus differentiated service delivery models, and TPT outcomes for people living with HIV in two of those countries, South Africa and Zimbabwe, where DSD models were operating at the study sites. We excluded the Opt4TPT data from Ethiopia because only one eligible participant received ART from a DSD model, which was insufficient for our analysis.

Patients in these countries receive ART through a variety of different HIV care models, including differentiated service delivery (DSD) models and the conventional clinic-based care model.

According to national guidelines, adults with undetectable viral loads (<50 copies/ml) on the same ART regimen for at least six (Zimbabwe) or 12 (South Africa) months are eligible to receive ART through a DSD model (24,25). Conventional care involves in-person visits with clinicians every one to two months, while DSD models involve an in-person visit with a clinician every 6- 12 months (4).

There were several DSD models in use in South Africa and Zimbabwe, including 1) fast-track ART pickup, where participants pick up their medication from a clinic pharmacy without interacting with a clinician; 2) family pick-up, where a family member can pick up their ART prescription from the fast-track pickup instead of the patient themselves; 3) community pick-up, where participants obtain their ART prescription from dispersion points in their community; and 4) community adherence groups, where DSD leaders in the community meet with patients to disseminate ART and discuss their ART adherence. The majority of HIV patients in these countries receive their ART from conventional models, though participation in DSD models is growing (10).

We enrolled participants in six public primary care clinics across two countries between August 17^th^ and December 29^th^ 2021. Health facility staff referred all PLHIV attending a clinic visit for ART initiation or prescription refills for study screening. Participants were eligible to participate and receive TPT if they met the following criteria: (a) age ≥ 18 years, (b) living with HIV, (c) initiating or refilling ART, (d) able and willing to attend in-clinic visits 12 and 24 months after enrollment. Participants were ineligible for TPT if they had previously taken TPT, by either self-report or according to their medical record.

### Procedures and Measures

#### Predictors

At the baseline visit, we collected demographic information on participants and the ART delivery model. We abstracted information about the participant’s clinical history, including the time they initiated ART and prior history of TB and TPT, from their clinical records. We repeated the survey on the ART delivery model at the participant’s six-month and twelve-month in-person appointments. As our primary predictor, we classified whether the participant received ART from a DSD model during their time on TPT or, if they never initiated TPT, if they received ART from a DSD model at their enrollment visit.

#### Outcomes

Guided by the RE-AIM (reach, effectiveness, adoption, implementation, maintenance) framework, we evaluated two key implementation outcomes: the reach of TPT, measured by the proportion of PLHIV who initiated TPT, and fidelity of TPT delivery, assessed by the implementation outcomes time to TPT initiation and the proportion of participants who completed treatment (23). If clinic nurses initiated TPT between the participant’s baseline visit and 12 months later, study nurses abstracted the start date from the medical records for inclusion in the study record. We defined initiation as beginning any TPT regimen within 12 months of study enrollment. We defined time to initiation as the number of days between the participant’s enrollment in the study and the day they initiated TPT.

For a proportion of participants, when they received TPT, it was dispensed in an EvriMED1000 MERM (Wisepill Technologies, Cape Town, South Africa), with reminders disabled. MERMs were given to consecutive participants until all boxes, based on the study budget, were used. We used the MERM to assess the TPT completion outcome. We defined completion for 3HP as taking ≥11 3HP doses within 16 weeks, for 6H (six months of daily Isoniazid) as completing ≥145/182 doses over 32 weeks, and for 12H (12 months of daily Isoniazid) as completing 292/365 doses over 64 weeks. We followed participants from their enrollment in the study to the end of their participation 24 months later.

Loss-to-follow-up was determined if participants did not attend their 12-month or subsequent follow-up visits.

### Statistical Analysis

We summarized the participants’ baseline demographic and clinical characteristics using proportions for categorical variables and medians and interquartile ranges for continuous variables. We used the chi-squared test for categorical variables and the independent samples t-tests for continuous variables to compare the characteristics of the participants receiving ART from DSD and the conventional care groups.

We constructed multivariate logistic regression models to assess the association of receiving ART from a DSD model with TPT initiation and TPT completion, adjusting for previously reported potential confounders, including sex, age (26), country, smoking, years on ART, and previous TB treatment. For the TPT completion model, we adjusted for the additional potential confounders TPT regimen, marriage, and employment in the TPT completion model.

For the TPT completion model, we used complete case analysis because the MERM boxes were only given to a portion of the participants. We conducted a sensitivity analysis for the TPT completion outcome using the multiple imputations by chained equations (MICE) method to address potential biases introduced by missing data. We used this approach because the data was assumed to be missing at random (MAR). This means the probability of missingness is related to observed data but not the missing data itself, based on observed predictors. We assessed collinearity using Pearson’s correlation, goodness-of-fit using the Hosmer-Lemeshow test, and model stability using ordinary nonparametric bootstrapping. We examined 95% confidence intervals in lieu of power calculations, given the fixed sample size.

We used the log-rank test and a Cox proportional hazards model, adjusting for the same potential confounders as the TPT initiation model (including sex, age, country, smoking, years on ART, and previous TB treatment), to compare the time to treatment initiation between people receiving HIV from DSD and conventional models. Participants who did not initiate TPT by the end of the initiation follow-up period (12 months or their last follow-up visit in the first 12 months if they were lost to follow-up) were right-censored. We addressed the clustering of outcomes at specific time points with the Efron tie method, which adjusts the likelihood function to distribute risk evenly over all participants who initiated at those clustered times. We analyzed all data using R (RStudio, Boston, MA) (27).

### Ethics Statement

The study protocol was approved by the University of the Witwatersrand Human Research Ethics Committee (#210212), the Medical Research Council of Zimbabwe (#MRCZ/A/2727), and the Yale University Human Investigation Committee (#2000035520). Formal written consent was obtained from the participants.

## Results

### Characteristics of the study population

Of the 1509 PLHIV screened for this study, 1193 were enrolled. Of the people screened, 237 were ineligible, including 193 who had previously taken TPT, 34 who were not willing to attend in-clinic visits at 12 and 24 months, four who were <18 years old, four who were not taking ART, and two who were not diagnosed with HIV. Of the people screened, 79 did not consent to participate (Figure 1).

**Fig 1.**
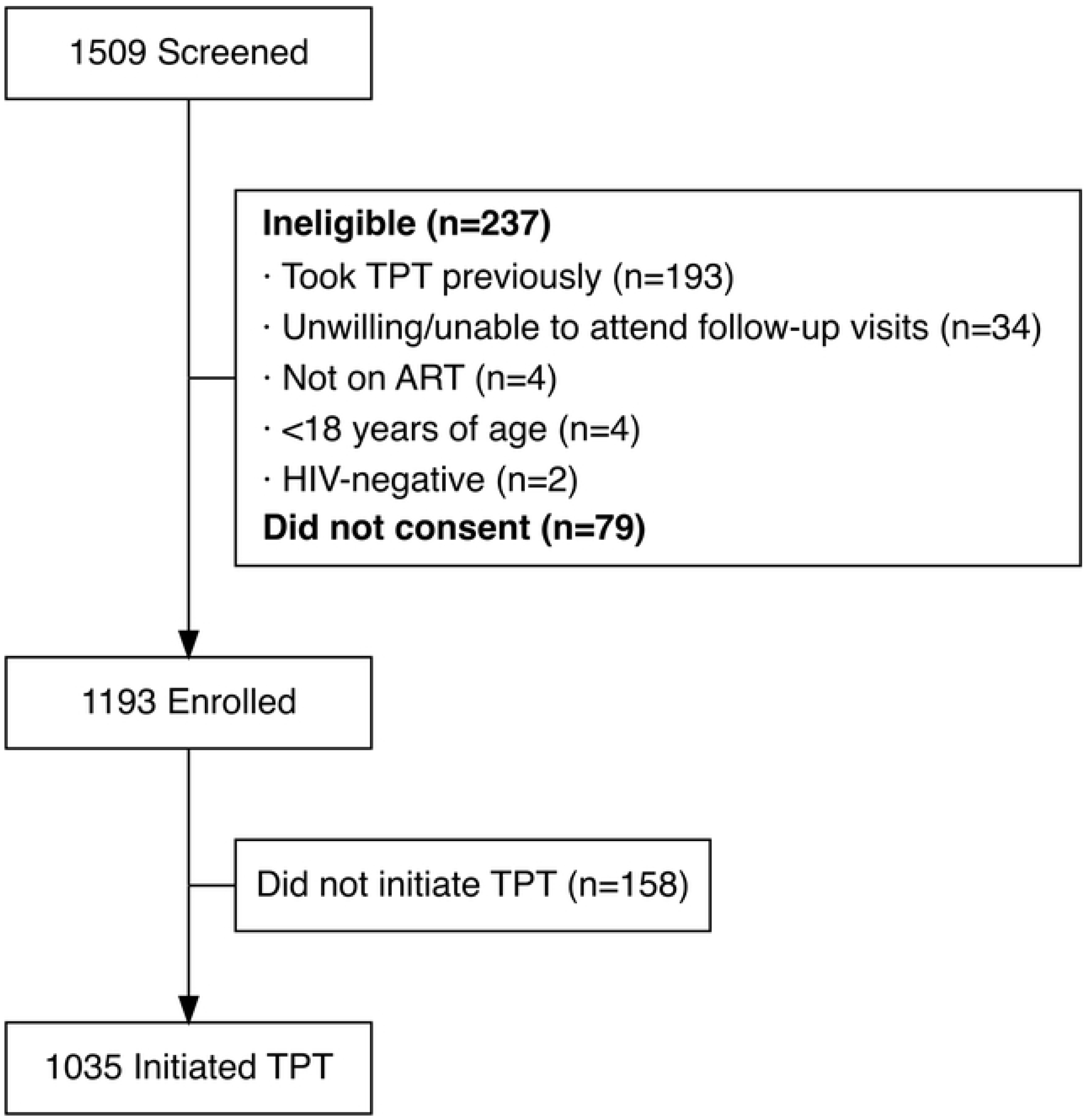
Enrollment Flow Diagram

The participants included 830 (70%) women, with a median age of 42 (interquartile range, IQR 15). There were 585 participants in South Africa and 608 in Zimbabwe. There were 276 (23%) participants in the DSD models, including 217 in South Africa and 59 in Zimbabwe. In the conventional care model, there were 917 (77%) participants. The types of DSD models participants were using included clinic fast lane pickup (n=182), community adherence groups (n=46), community pickup (n=44), and family pickup (n=4).

There were equal numbers of men and women in the DSD and conventional care models. However, in the DSD group, there were higher proportions of participants with viral loads below 50 copies/mL. In contrast, the conventional group had a higher proportion of participants with viral loads in the 500+ copies/mL range (Table 1). In the DSD group, participants had a median time on ART of 8 years (IQR 7), while the conventional group had a shorter median time on ART of 6 years (IQR 9, Table 1). There were significantly (p < 0.001) more participants in the DSD group who received 3HP, while the conventional group had a higher proportion of participants with the 6H regimen.

**TABLE 1.** Characteristics of PLHIV Receiving ART, by Delivery Model.

| Characteristic n=1193 | Conventional n=917 | DSD n=276 | *p* value |
| --- | --- | --- | --- |
| Age (years) | 41 (15) | 45 (12) | <0.001<sup>3</sup> |
| Female Sex | 634 (69%) | 196 (71%) | 0.60<sup>4</sup> |
| Country |  |  | <0.001<sup>4</sup> |
| South Africa | 368 (40%) | 217 (79%) |  |
| Zimbabwe | 549 (60%) | 59 (21%) |  |
| Married | 442 (48%) | 103 (37%) | 0.002<sup>4</sup> |
| Employed | 191 (21%) | 51 (18%) | 0.44<sup>4</sup> |
| Smokes | 160 (17%) | 68 (25%) | 0.010<sup>4</sup> |
| Prior treatment for active TB | 96 (10%) | 27 (10%) | 0.83<sup>4</sup> |
| Diabetic | 22 (2%) | 5 (2%) | 0.73<sup>4</sup> |
| Time on ART (years) | 6 (9) | 8 (7) | <0.001<sup>3</sup> |
| Viral load (copies/mL)<sup>1</sup> |  |  | 0.001<sup>4</sup> |
| <50 | 619 (68%) | 208 (75%) |  |
| 50-499 | 90 (10%) | 49 (18%) |  |
| ≥500 | 45 (5%) | 5 (2%) |  |
| 3HP Regimen<sup>2</sup> | 387 (42%) | 184 (67%) | <0.001<sup>4</sup> |

**Abbreviations:** ART, Antiretroviral Therapy; 3HP, 3-month once-weekly isoniazid-rifapentine; DSD, Differentiated Service Delivery; IQR, Interquartile Range; TB, Tuberculosis; TPT, Tuberculosis Preventive Therapy
**Legend:** <sup>1</sup>Missing: 177; <sup>2</sup>For those who initiated, n=1035, Missing: 16; Other Regimens: 12H n=4, 6H n=444; <sup>3</sup>T-test; <sup>4</sup>Chi-squared test

### Reach

In total, 1035 (87%) of the 1193 participants initiated TPT over the initial 12 months of the study. 242 (88%) of the 276 participants receiving ART from a DSD model initiated TPT, and 793 (86%) of the 917 participants receiving ART from a conventional model initiated TPT (Table 2A). The initiation proportion was similar in community pickup (82%) and clinic fast pickup (86%) models, but it was 100% in the community adherence group (n=46) and family pickup models (n=4).

**TABLE 2A.** TPT Outcomes Across ART Delivery Models.

| Outcome | DSD n=276 | Conventional<br>n=917 | OR (95% CI) | p value | Adjusted OR (95% CI) | Adjusted p<br>value |
| --- | --- | --- | --- | --- | --- | --- |
| TPT Initiation | 242 (88%) | 793 (86%) | 1.11 (0.74-1.67) | 0.61 | 1.03 (0.66-1.61)<sup>1</sup> | 0.88<sup>1</sup> |
| TPT Completion | 82 (57%) | 274 (47%) | 1.53 (1.06-2.21) | 0.024 | 0.89 (0.58-1.36)<sup>2</sup> | 0.58<sup>2</sup> |

**Abbreviations:** DSD, Differentiated Service Delivery; TPT, Tuberculosis Preventive Therapy
**Legend:** <sup>1</sup>Adjusted for sex, age, marriage, employment, country, smoking, years on ART and previous TB treatment; <sup>2</sup>Adjusted for sex, age, marriage, employment, country, smoking, years on ART, previous TB treatment and TPT regimen

In bivariate analysis, the odds of initiating TPT were significantly higher among those who were older (OR 1.23, 95% CI 1.06-1.44, p = 0.007) and who had been receiving ART for longer (OR 1.04, 95% CI 1.00-1.08, p = 0.030). We did not find a significant association between receiving ART from a DSD model (OR 1.11, 95% CI 0.74-1.67, p = 0.61) and TPT initiation. Sex, marriage, country, employment status, previous TB treatment, and smoking were also not significantly associated with TPT initiation.

In multivariate analysis, we did not find a significant association between receiving ART from a DSD model and initiating TPT (OR 1.03, 95% CI 0.66-1.61, p = 0.88) after adjusting for the potential confounders sex, age, marriage, employment, country, smoking, years on ART and previous TB treatment.

### Implementation

DSD models had, on average, a longer average time to initiation of 6.5 days (standard deviation (SD) = 34.6) compared to conventional models (mean 2.7 days, SD = 18.7, Table 2B). Most participants who initiated TPT throughout the study did so at their enrollment visit, including 645 (81%) of those in conventional models and 124 (51%) of those in DSD models, so both groups had a median time to initiation of 0.

**TABLE 2B.**
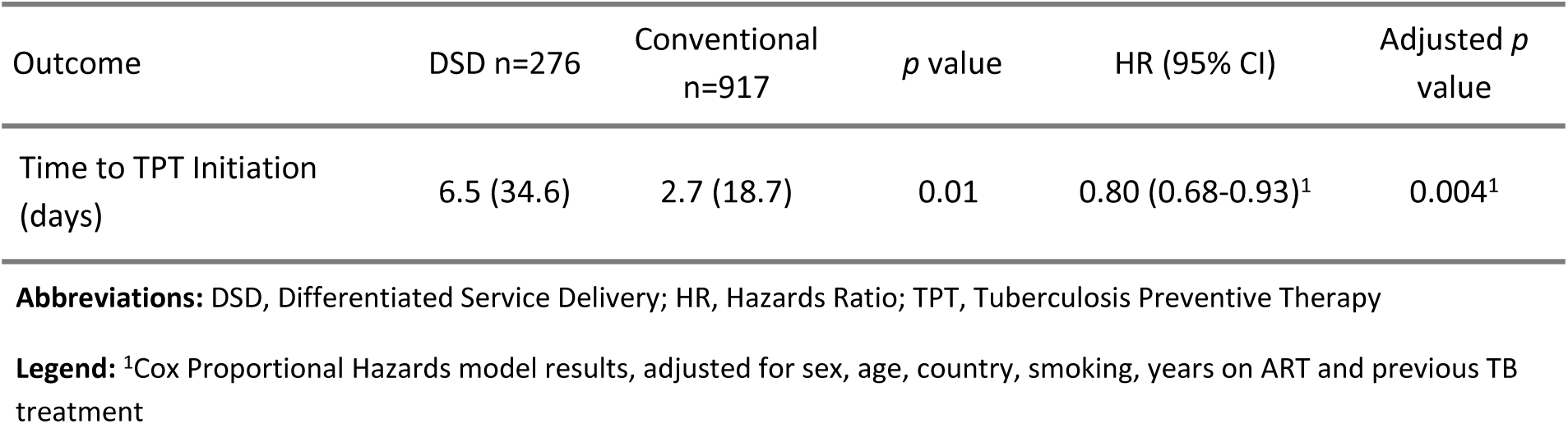
TPT Outcomes Across ART Delivery Models.

| Outcome | DSD n=276 | Conventional<br>n=917 | *p* value | HR (95% CI) | Adjusted *p*<br>value |
| --- | --- | --- | --- | --- | --- |
| Time to TPT Initiation<br>(days) | 6.5 (34.6) | 2.7 (18.7) | 0.01 | 0.80 (0.68-0.93)<sup>1</sup> | 0.004<sup>1</sup> |

**Abbreviations:** DSD, Differentiated Service Delivery; HR, Hazards Ratio; TPT, Tuberculosis Preventive Therapy
**Legend:** <sup>1</sup>Cox Proportional Hazards model results, adjusted for sex, age, country, smoking, years on ART and previous TB treatment

The log-rank test showed a significant difference in time to TPT initiation, with participants receiving ART from DSD models having a statistically significantly longer time to treatment initiation (p= 0.01) than participants receiving ART from conventional models. In the Cox proportional hazards model, after adjusting for sex, age, country, smoking, years on ART, and previous TB treatment, receiving ART from DSD models was significantly associated with a 20% lower hazard of initiating TPT at any given time point (HR = 0.80, 95% CI 0.68-0.93, p = 0.004). As is shown in the hazards curve (Figure 2), there is initially a minimal initiation difference between the groups. However, TB incidence rates per day are low enough that it’s not likely an important difference in time to initiation (28).

**Fig 2.**
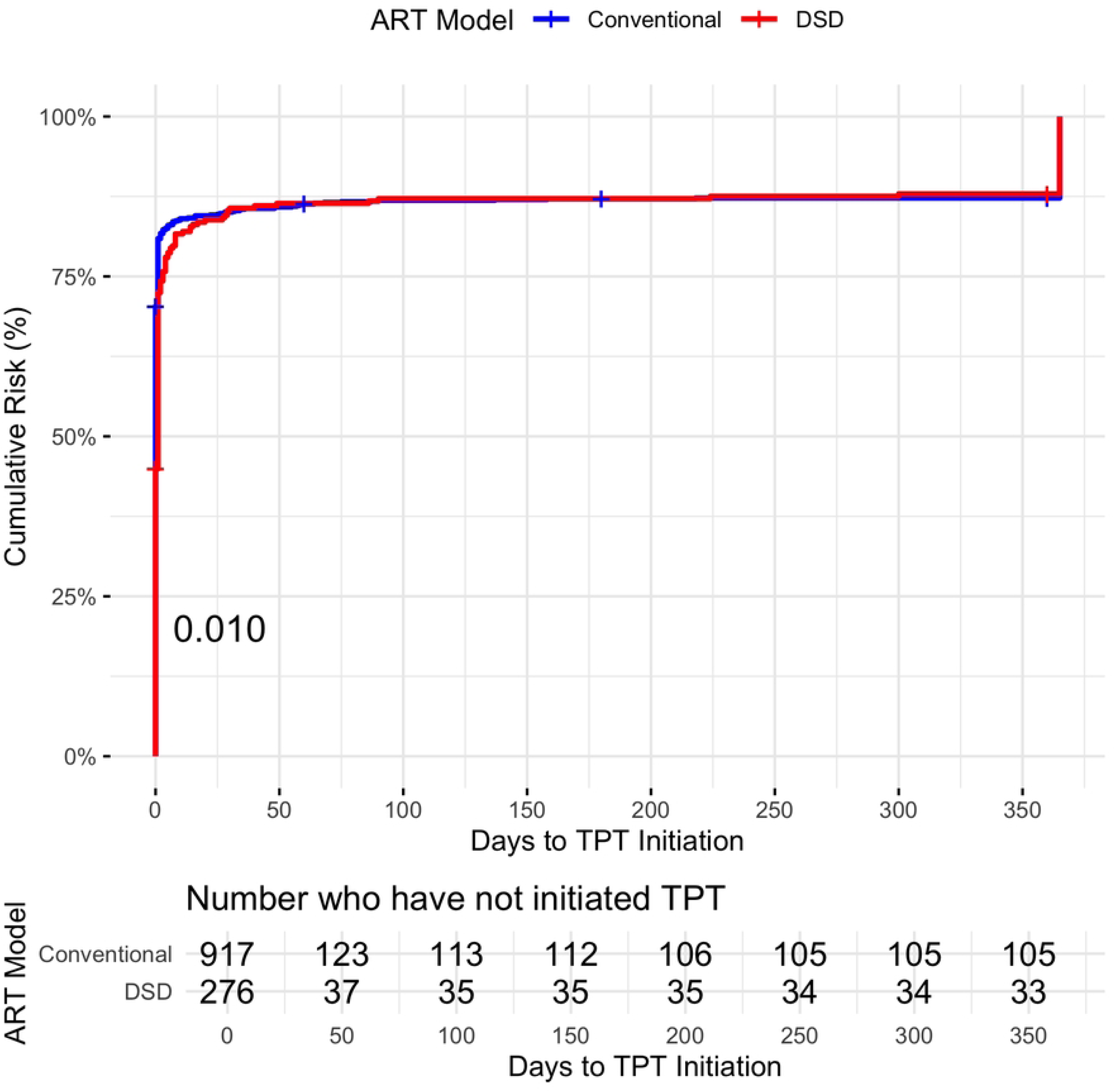
Hazard Curve of Time to TPT Initiation for DSD vs. Conventional Models

We have completion data for 731 (71%) of the 1035 participants who initiated TPT. Of these, 356 (49%) completed TPT. Of the participants receiving ART from a DSD model, 82 (57%) completed TPT, and 274 (47%) received ART from a conventional model completed TPT.

The completion proportion varied from 11% in community adherence groups to 100% with family pickup, though the small numbers in these groups limit the precision of these estimates.

In bivariate analysis, the odds of completing TPT were significantly higher among participants who receive ART from DSD models (OR 1.53, 95% CI 1.06-2.21, p=0.024) and who received 3HP (OR 2.94, 95% CI 2.15-4.01, p <0.001). In multivariate analysis, we did not find a significant association between receiving ART from a DSD model and completing TPT (OR 0.89, 95% CI 0.58-1.36, p=0.58) after adjusting for sex, age, marriage, employment, country, smoking, years on ART, previous TB treatment and 3HP regimen (Table 2A).

The Pearson correlation coefficients indicated moderate correlations between several predictors in the multivariate completion analysis. In particular, receiving 3HP was moderately correlated with receiving ART from a DSD model (PCC= 0.33) and country (PCC= 0.42).

### Sensitivity Analysis

22 (1.5%) of the participants did not initiate TPT and were lost to follow-up before their 12- month follow-up visit. Overall, out of the 1193 participants, 106 (9%) changed from a conventional model to a DSD model or a DSD model to a conventional model during the 24 months the participants were followed. After starting TPT, 62 changed from a conventional to a DSD model, and 10 changed from a DSD a conventional model.

Participants who received the 3HP regimen were more likely (p < 0.001) to have received the MERM box to measure completion. This suggests that the completion data is missing at random (MAR) because the probability of missing data is related to a measured variable, the TPT regimen. The Hosmer-Lemeshow test indicated improved goodness-of-fit in the imputed model (p = 0.72) compared to the complete case model (p = 0.16), and bootstrapping analyses confirmed the stability of model coefficients, indicating robust estimation under multiple imputation. These analyses suggest that the missing data did not significantly bias our results.

The multivariate logistic regression results based on the imputed data had the same significant associations between the potential confounders and TPT completion as the complete case analysis did with similar odds ratios and confidence intervals, and the results were therefore not sensitive to the choice of analytic approach.

## Discussion

In this longitudinal cohort study, there was a high TPT initiation rate in PLHIV in DSD and conventional models, showing that receiving ART from a DSD model was not associated with lower TPT initiation. Receiving ART from DSD models was significantly associated with a lower hazard of initiating TPT at any given time. However, the actual difference in time to initiation was small and likely not meaningful. Completion rates were low overall, and DSD models had a higher completion rate than conventional models. We did not find a significant independent association between receiving ART from DSD models and completing TPT. The high TPT initiation rates in DSD models demonstrate that distributing TPT to people receiving care from DSD models is feasible. However, the low completion rates across both models indicate TPT completion as an important area of focus for improving TPT delivery, which is essential because it is not just initiating but completing TPT that significantly reduces TB deaths in people with HIV (29).

The analyses of the association of receiving ART from a DSD model on TPT initiation and TPT completion were underpowered to detect a true difference, according to the 95% CI for each analysis (30). The 95% confidence intervals indicate that the true effect could range from a 34% decrease to a 61% increase in the odds of initiating TPT and from a 42% decrease to a 36% increase in the odds of completing TPT. This study should be considered exploratory, and future research with a larger sample of participants in DSD models should be conducted to accurately determine the effect of receiving HIV from DSD models on initiating TPT, as a higher-powered study is necessary to capture the potential impact outlined by this confidence interval reliably.

Reports of TPT initiation rates in DSD models are limited and varied. In Uganda, an intervention aligned TPT refills with ART refills and expanded DSD model capacity, which increased TPT initiation from 22% to about 60% (31). In contrast, a 2021 study of South African and Eswatini DSD models reported only a 4% rate of Isoniazid preventive therapy for PLHIV, which they attributed to the possibility that many PLHIV had already taken TPT (17). Our study found a higher initiation rate, potentially due to the availability of the 3HP regimen and because the presence of the study analyzing TPT at the clinics probably increased TPT enrollment rates.

Introduced in 2021, 3HP can be given as a single 3-month course in one dispensing. This may be more compatible with the lower frequency of clinic visits in DSD models and could have contributed to the higher initiation rates observed in our study (32).

Several studies have examined TPT completion for PLHIV in DSD models, including those conducted in 2017 (26) and 2019 (33) in Uganda and a 2023 study in Zambia (34). All three studies measured completion only with the 6H regimen (6 months of Isoniazid). The Ugandan studies reported completion rates around 70 and 90%, and the Zambian study found around 90% completion. These studies measured completion by patients picking up their final month refill, self-report, or clinician report. These methods tend to overestimate completion compared to the objective MERM box measure used in our study (35,36). Our study’s lower completion rates may be attributed to these differences in measurement methods.

Though there was a significant difference in time to initiation between the groups, the actual difference between mean time to initiation is small (3.8 days). The incidence of tuberculosis for people with well-controlled HIV (viral load <50), like the majority of participants in this study, are likely to have higher CD4 counts and lower TB incidence (28), so there is likely no meaningful difference between these times to initiation. DSD models may have had a longer time to initiation because participants in those models have fewer visits with their healthcare providers, so they may have less time with providers to focus on preventive healthcare such as TPT.

A higher proportion of participants in the DSD models received their TPT in the form of 3HP, potentially because the single disbursement of 3 months of pills meant the participants with this regimen did not have to attend extra visits outside their normal DSD visits or because participants in DSD models are more likely to be eligible for 3HP than participants in conventional models. People receiving 3HP were much more likely to complete treatment compared to people on other TPT regimens, as in previous studies (37,38). So, the 3HP regimen could be a confounding variable that explains why the difference in TPT completion in DSD and conventional models was significant in bivariate analysis but not independently after the adjustment for regimen and the other potential confounders.

The low overall TPT completion rate indicates the need for innovations to support TPT completion. Supportive measures for providing TPT through DSD models have been trialed for the 6H regimen, such as multi-month dispensing of 6H to better align with DSD appointment schedules, reminder phone calls during the treatment period, and structured patient education on TPT (34). A similar adaption for providing preventive treatment through DSD models was the scale-up of DSD-based multi-month dispensing of Pre-exposure prophylaxis (PrEP) during COVID-19, which significantly increased PrEP uptake (39). Data should be collected on the impact of similar supportive measures for 3HP on objective completion measures. Also, data on uptake and adherence to preventive and noncommunicable disease treatment should be collected to understand these groups better as DSD’s reach expands.

Our study had several strengths. We controlled for potential confounders such as ART history and previous TB treatment by collecting comprehensive baseline data, which strengthened the reliability of our findings. We also had objective measures for our outcomes, including clinical abstracts of TPT initiation and MERM data to measure TPT completion. This was an observational study of TPT initiation and completion during the TPT scale-up in Zimbabwe and South Africa, so the results may be generalizable for the scale-up of treatment in other settings. We included participants from two countries, South Africa and Zimbabwe, and six primary care clinics, which enhances the generalizability of our results to similar contexts in sub-Saharan Africa. We also included participants from four different types of DSD models, which improves the generalizability of the findings to DSD models broadly.

Our study also had several limitations typical of longitudinal cohort studies. Patients in conventional models had visits every three months, and patients in DSD models had clinic visits every 6-12 months, which resulted in a higher representation of conventional model participants. Due to this, the study was underpowered, and we were unable to examine the different DSD models separately. TPT was delivered during visits at the clinics, so delivering TPT through community settings may yield different results. We measured completion objectively with the MERM, though completion could be overestimated if participants opened the boxes without taking the medication or underestimated if they used medications obtained from other sources. Additionally, the ease of using MERM boxes might have inadvertently boosted completion, even with the reminders disabled. Missing data was a potential limitation, but our analysis suggests this is unlikely to have influenced our results. Because this data was collected during the TPT scale-up, the high initiation rates and short times to initiation may not be generalizable to routine, past initial scale-up clinical environments.

We did not find major differences between TPT initiation or completion between DSD and conventional models. The results are encouraging that TPT delivery in DSD models is feasible. However, the robustness of the results may be limited by the small sample size of participants in DSD models. TPT completion was low across both models, so increasing completion should be a focus of future research. Future large prospective and pragmatic studies should address the integration of TPT and other lifesaving preventive interventions for communicable and non-communicable diseases into DSD models in overburdened healthcare systems. Adapting DSD models with support specific to TPT, such as aligning distribution appointments and providing reminder phone calls, could increase TPT initiation and completion in sub-Saharan Africa.

## Data Availability

All relevant data underlying the findings of this study are provided within the paper and its Supporting Information files. No additional data are required to replicate the analyses described in this manuscript.

## Acknowledgements

Sources of Funding: This publication was made possible by CTSA Grant Number TL1 TR001864 from the National Center for Advancing Translational Science (NCATS), a component of the National Institutes of Health (NIH). Its contents are solely the responsibility of the authors and do not necessarily represent the official views of NIH.

This study was supported by the Bill and Melinda Gates Foundation through Investment number INV-006096.

AEJ was supported by NIH Medical Scientist Training Program T32 GM136651 and the Global Health Equity Scholars (GHES) Program, which is sponsored by the Fogarty International Center (FIC) and several collaborating Institutes and Centers at the National Institutes of Health (NIH).

## Notes

### Competing Interest Statement

The authors have declared no competing interest.

### Funding Statement

No authors have financial disclosures.

### Author Declarations

The study protocol was approved by the University of the Witwatersrand Human Research Ethics Committee (#210212), the Medical Research Council of Zimbabwe (#MRCZ/A/2727), and the Yale University Human Investigation Committee (#2000035520).

